# Effect of High-Frequency Repetitive Transcranial Magnetic Stimulation on Upper Limb Function After Stroke: A Systematic Review and Meta-Analysis

**DOI:** 10.1101/2025.01.23.25320584

**Authors:** Nazzareno Russo, Thomas Hunt, Alex McMullin, Timothy Payard, Elise Gane, Martin Sale

## Abstract

Functional recovery of motor deficits post-stroke is improved with physical therapy, however functional recovery is often incomplete. Non-invasive brain stimulation may enhance functional recovery post-stroke. This systematic review explored the effectiveness of high-frequency repetitive Transcranial Magnetic Stimulation (HF-rTMS) to improve upper limb motor function post-stroke. A database search was conducted within PubMed, CINAHL, Embase and Cochrane Library. Criteria for inclusion: randomised controlled trials employing HF-rTMS on the affected hemisphere of adults post-stroke, and using the Fugl-Meyer Assessment (FMA) tool. The Cochrane Risk of Bias 2 tool and GRADE framework were applied. Sixteen articles were included in the review. There was significant improvement in FMA-UL scores immediately post-intervention (mean difference = 3.53 [95% CI 1.82 to 44.58]) and 1-3 months post-intervention (8.95 [5.95 to 11.95]) for patients who commenced treatment within 1-month of their stroke. Based on the FMA, excitatory rTMS may provide more favourable effects on motor recovery when applied in the first-month post-stroke, however a variety of heterogeneous application parameters limit the certainty of effectiveness.

## 1. Introduction

Cerebrovascular accidents (CVAs) or strokes are categorised as either haemorrhagic or ischemic in origin. Both, however, lead to compromise of cerebral perfusion and ultimately hypoxia-induced cell death. Immediately following a stroke, the affected cortical regions can lose 4 million neurons, and roughly 15 billion synapses every minute if left untreated (Hassan & Rohatgi, 2009). As a result, the capacity of these areas to operate functionally is significantly impaired. Accordingly, ischemic insults to the motor cortex can present with altered or absent neuromuscular related functions. Persistent upper limb weakness or hemiparesis is not only a common contributor to the reduced quality of life experienced by stroke victims but also by their caregivers (Lai et al., 2002; Opara & Jaracz, 2010; Raghavan, 2015). Current statistics indicate that 1 in 4 people will experience a stroke within their lifetime (Feigin et al., 2022). Furthermore, with over 143 million years of healthy life lost annually as a result of stroke-related death and disability, the estimated global cost exceeds US $720 billion (Feigin et al., 2022).

As a result of ischemic lesions in the primary motor cortex (M1), ongoing functional impairments can, in part, be attributed to altered cortical excitability in surrounding areas (Ward & Cohen, 2004). Functional recovery following a stroke relies on adjacent cortical regions to compensate for the damaged motor areas through somatotopic reorganisation (Beaulieu & Milot, 2018). This is highly dependent on neuroplasticity. These neuroplastic changes are promoted by large doses of physical rehabilitation that focus on performing and re-learning the affected motor tasks (Khan et al., 2019). However, a significant proportion of stroke survivors will suffer from persistent impairments in upper limb function despite large doses of contemporary rehabilitation (Lai et al., 2002; Raghavan, 2015). By modulating cortical excitability, emerging forms of non-invasive brain stimulation (NIBS) techniques may promote neuroplastic changes in the underlying cortex. One common form of NIBS used in stroke rehabilitation is Transcranial Magnetic Stimulation (TMS).

TMS produces a transient and focal magnetic field above the skull, inducing an electrical stimulus in the area of the brain directly beneath the coil which can ultimately activate peripheral muscles controlled by the stimulated cortical region (Burke et al., 2019; Klomjai et al., 2015; van Lieshout et al., 2019). The painless, non-invasive application of TMS is well tolerated by participants and provides similar effects to that of direct electrical stimulation to the surface of the brain without the associated pain of electrical stimulation (Ridding & Rothwell, 2007). Although a single pulse of TMS can induce transient changes to cortical excitability, repetitive TMS (rTMS) can generate prolonged after-effects (Klomjai et al., 2015).

The application of rTMS to impaired cortical pathways is thought to induce long-lasting changes in function via its effect on synaptic plasticity (Ridding & Rothwell, 2007). Cortical neurons surrounding stroke-affected areas become hypoactive and thus are functioning sub-optimally. By using an excitatory paradigm, rTMS can increase excitability in these regions, and thus training-induced synaptic plasticity is more likely to occur (Kim et al., 2006; Stefan et al., 2000). Consequently, motor function may be enhanced by increasing cortical excitability with the application of excitatory rTMS to ipsilesional brain regions (Bäumer et al., 2003; Ridding & Rothwell, 2007). rTMS can be delivered at different frequencies. High-frequency (HF) stimulation is delivered to ipsilesional regions and utilises processes of long-term potentiation (LTP) to increase the excitability of the neurons in the targeted area (Stefan et al., 2006; Ziemann et al., 2004). In contrast, low-frequency (LF) stimulation is delivered to contralesional regions and utilises processes of long-term depression (LTD) to decrease the excitability of the neurons in the targeted area (Stefan et al., 2006; Ziemann et al., 2004). Consistent excitation (or depression) of synaptic signalling can lead to long-lasting changes in neuronal activity (Ridding & Rothwell, 2007). Low-frequency rTMS (LF-rTMS) has been more widely studied for its potential therapeutic benefits in stroke patients, particularly in reducing spasticity and improving motor function (Fisicaro et al., 2019; Sharma et al., 2020; Yao et al., 2020; Zhang, Xing, Shuai, et al., 2017).

The majority of previous meta-analyses have investigated the utility of inhibitory (<1Hz) or low-frequency rTMS (LF-rTMS) (Fisicaro et al., 2019; Sharma et al., 2020; Yao et al., 2020; Zhang, Xing, Shuai, et al., 2017). As a result, there is limited evidence for the effectiveness of excitatory (>1Hz) or high-frequency rTMS (HF-rTMS) in acute and chronic stroke patients (Graef et al., 2016; Khan et al., 2019; van Lieshout et al., 2019). Limited commonly by small sample sizing and between-study heterogeneity of outcome measures, some tentative evidence suggests rTMS may induce short- and long-term therapeutic effects on upper limb motor function after stroke (van Lieshout et al., 2019; Zhang, Xing, Fan, et al., 2017). Research indicates early intervention of neurorehabilitation maximises clinically significant functional effects (Alia et al., 2017; Kleim & Jones, 2008; Zeiler & Krakauer, 2013). Meaningful functional recovery has been reported in physiotherapy interventions that are introduced within 1-3 months post-stroke; this time frame coincides with the greatest capacity for neuroplastic changes (Alia et al., 2017; Kleim & Jones, 2008; Zeiler & Krakauer, 2013). Currently, however, there is inconclusive evidence on rTMS application parameters (e.g., stimulation frequency, number of pulses delivered, intervals of rTMS application, and the number of sessions involved), as well as the timing of initial treatment with rTMS in acute, subacute, and chronic phases of stroke (Bernhardt et al., 2015).

Inconsistent outcome measures have also limited the ability to draw firm conclusions regarding the efficacy of rTMS on upper limb motor function (Beaulieu & Milot, 2018; Vabalaite et al., 2021). Largely due to limited data, previous meta-analyses and systematic reviews have been unable to account for the variation in functional outcome measures of the upper limb (van Lieshout et al., 2019; Zhang, Xing, Fan, et al., 2017). The International Classification of Function, Disability and Health (ICF) model encompasses outcome measures at the level of function, activity, or participation (World Health, 2001). Improvements in upper limb function due to rTMS are more likely to be detected using functional outcome measures such as the Fugl-Meyer Arm (FMA) assessment, and a stroke-specific motor recovery test (van Lieshout et al., 2019). Previous literature has regarded function as a more reliable outcome measure for distinguishing changes at the neural level compared with activity and participation outcome measures, which are influenced greatly by cognitive, personal and environmental factors (Gladstone et al., 2002; van Lieshout et al., 2019).

Having a greater understanding of the efficacy of HF-rTMS as a rehabilitation tool post-stroke may have significant implications for functional recovery. With this in mind, we conducted a systematic review with the aim to explore the effectiveness of HF-rTMS in improving upper limb motor function in patients post-stroke.

## 2. Methods

This systematic review was conducted in accordance with the Preferred Reporting Items for Systematic Reviews and Meta-Analyses (PRISMA) statement (Page et al., 2021). This review was not registered with PROSPERO because it was conducted within the context of a postgraduate course at a university, and PROSPERO does not accept the registration of reviews done as part of training courses.

### 2.1 Search Strategy

Four reviewers (AM, TP, NR and TH) conducted a comprehensive search strategy on published literature within PubMed, CINAHL, Embase and Cochrane Library databases. Within these databases, randomised trials, full-text and English language limits were applied as able. Search strategies were informed by the population of interest (P), the intervention (I), the comparator (C), and the outcome (O), otherwise known as the PICO framework. More specifically, each search strategy contained four key search strings; namely transcranial magnetic stimulation, stroke, upper limb, and motor function. To increase the scope of the search strategy, the outcome of interest was separated into two strings, upper limb and motor function respectively. Search terms were applied to both subject headings and text-based field codes. A full outline of the search strategy can be found in Appendix A.

### 2.2 Eligibility Criteria

Eligibility criteria were developed based on accepted definitions and established parameters sourced from previous studies. Studies were eligible for inclusion if they included; (1) patients were diagnosed with a stroke; (2) excitatory rTMS (>1 Hz) as the primary intervention studied; (3) the upper limb domain of the FMA as a primary or secondary outcome measure; (4) effects of excitatory rTMS were compared to a sham/control group; (5) a randomized controlled trial (RCT) study design; (6) patients >18 years of age; (7) written in English; (8) available full texts published in a peer-reviewed journal; and (9) published between 2010 and 2022.

### 2.3 Study Selection

References from each database search were imported to EndNote 20.2 and following the removal of duplicate files, the library was then uploaded to Covidence for screening. Two reviewers (AM and TP) evaluated each study based on title and abstract eligibility, while a further two reviewers (NR and TH) resolved conflicts. Full-text screening was conducted by four reviewers (AM, TP, NR and TH) with further conflicts resolved through discussions among all team members. Interrater reliability between all team members was assessed using the kappa statistic. Lastly, one reviewer (TP) conducted a secondary screening of reference lists and forward citation searching.

### 2.4 Data Extraction and Critical Appraisal of Studies

The following data were extracted from the included studies; study overview including aim, design, and setting; demographic and clinical characteristics of the participants including age, sex, time post-stroke, stroke location/type (if provided), the total number of participants per group, and FMA baseline score; intervention and control group characteristics including intervention and control rTMS protocol, additional therapy protocol, and quality of delivery; and finally, results of the intervention and control group as measured with the FMA. When there were missing data, authors of the published studies were contacted. Timing of treatment post-stroke was categorised following previous meta-analyses that utilised recommendations by the Stroke Recovery and Rehabilitation Roundtable (SRRR): acute to early-subacute (<1-month), early-subacute to late-subacute (1-6 months), and chronic (>6-months). The SRRR classification of acute (1-7 days) and early subacute (7 days-3 months) were combined and divided into acute to early-subacute (<1-month) and early subacute (1-3 months) to better represent any improved motor function within the first-month post-stroke, and to depict more accurately, the critical window of neuroplasticity. The early-subacute (1-3 months) and late-subacute (3-6 months) phases were also combined (1-6 months) in order to accommodate all included studies (Kwakkel et al., 2017).

Risk of bias of the included studies was evaluated using the Cochrane Risk of Bias tool (RoB2) for randomised controlled trials (Sterne et al., 2019). The tool is structured into five domains; (1) bias arising from the randomization process; (2) bias due to deviations from intended interventions; (3) bias due to missing outcome data; (4) bias in the measurement of the outcome; and (5) bias in the selection of the reported result (Sterne et al., 2019). Following responses to signalling questions, each domain received a risk of bias judgement (low, high, or some concerns) followed by an overall risk of bias judgement across all domains (low, high, or some concerns). Four reviewers (AM, TP, NR and TH) were involved in risk of bias assessment. Each study was independently evaluated by two different reviewers from the four included, conflicts were resolved through discussions among all team members.

The certainty of evidence across studies was assessed using the Grading of Recommendations, Assessment, Development and Evaluation (GRADE) framework. The framework features five domains: (1) risk of bias; (2) inconsistency; (3) indirectness; (4) imprecision; and (5) publication bias (Balshem et al., 2011). The certainty of evidence for the outcome of upper limb motor function was rated on a 4-point scale: very low, low, moderate or high.

### 2.5 Data Analysis

In order to facilitate a comparison between studies, effect sizes were calculated using Hedge’s g, with the addition of utilising the bias correctional factor (J) to account for studies with small sample sizes. Mean changes between baseline and post-intervention measurement in the HF-rTMS and control groups were divided by the pooled and sample-weighted standard deviation. Standard deviation was calculated when only the standard error of the mean was reported, or if the within-groups standard deviation between baseline and assessment was not disclosed. All calculations used were consistent with those available in the Cochrane Handbook that were appropriate relative to the available data reported per article. If no numerical data were reported, we extracted these from the figures, using PlotDigitizer 2.6.9 based on the Cochrane Handbook for Systematic Reviews of Intervention (Higgins et al., 2022). Consistent with previous systematic reviews and meta-analyses, where studies reported repeated outcome assessments, the first assessment performed after the treatment was used to represent the immediate post-intervention data (van Lieshout et al., 2019). Subsequent follow-up findings were then compared to baseline measures to calculate the effect size of each respective assessment. Utilizing the bias-corrected effect size (Hedge’s g), standardized mean differences (SMDs) with 95% confidence intervals (95% CIs) were used.

To prepare data for meta-analysis, studies were grouped by length of time since stroke at recruitment and time of assessment after the intervention. Five groups of studies were formed: (1) participants <1-month post-stroke, assessed immediately after intervention; (2) participants <1-month post-stroke, assessed 1-3 months after intervention; (3) participants <1-month post-stroke, assessed 6-12 months after intervention; (4) participants 1-6 months post-stroke, assessed immediately after intervention; and (5) participants >6-months post-stroke, assessed immediately after intervention. In groups of studies that cover a range of assessment time periods (groups 2-5), a study was only included once if results were available for more than one assessment time point in that study, and the longer time frame was chosen (e.g. 3 months was chosen over 1 month). This avoided a unit-of-analysis issue resulting from double counting. The GRADE framework was applied to each of these groups of studies.

Any form of HF rTMS was considered the intervention group, and any form of control or sham rTMS was considered the comparator group. The mean difference and SD for FMA-UL from each group as well as the sample size was imputed into RevMan (v5.4.1, The Cochrane Collaboration). Forrest plots were generated from models using fixed effects, inverse-variance methodology. The *I^2^* statistic was used to assess heterogeneity and interpreted as recommended by The Cochrane Collaboration: <40% might not be important; 30-60% moderate heterogeneity; 50-90% substantial heterogeneity, with 75-100% being considerable heterogeneity (Higgins et al., 2022)

## 3. Results

### 3.1 Study Selection

The database search produced 614 results, with 286 records screened at the title/abstract stage and 67 records at the full-text stage. After screening, 16 studies were deemed relevant and included in the review. Forward citation searching identified zero additional studies. Figure 1 outlines the screening results. When reviewing the title and abstracts, there was moderate to near-perfect agreement (range kappa 0.56 - 1.0) among the four reviewers (see Appendix B). When reviewing the full texts, most had a moderate to near-perfect agreement (range kappa 0.5 –1.0). However, TH and TP had only slight agreement on the 12 papers they both reviewed (range kappa 0.2).

**Figure 1.**
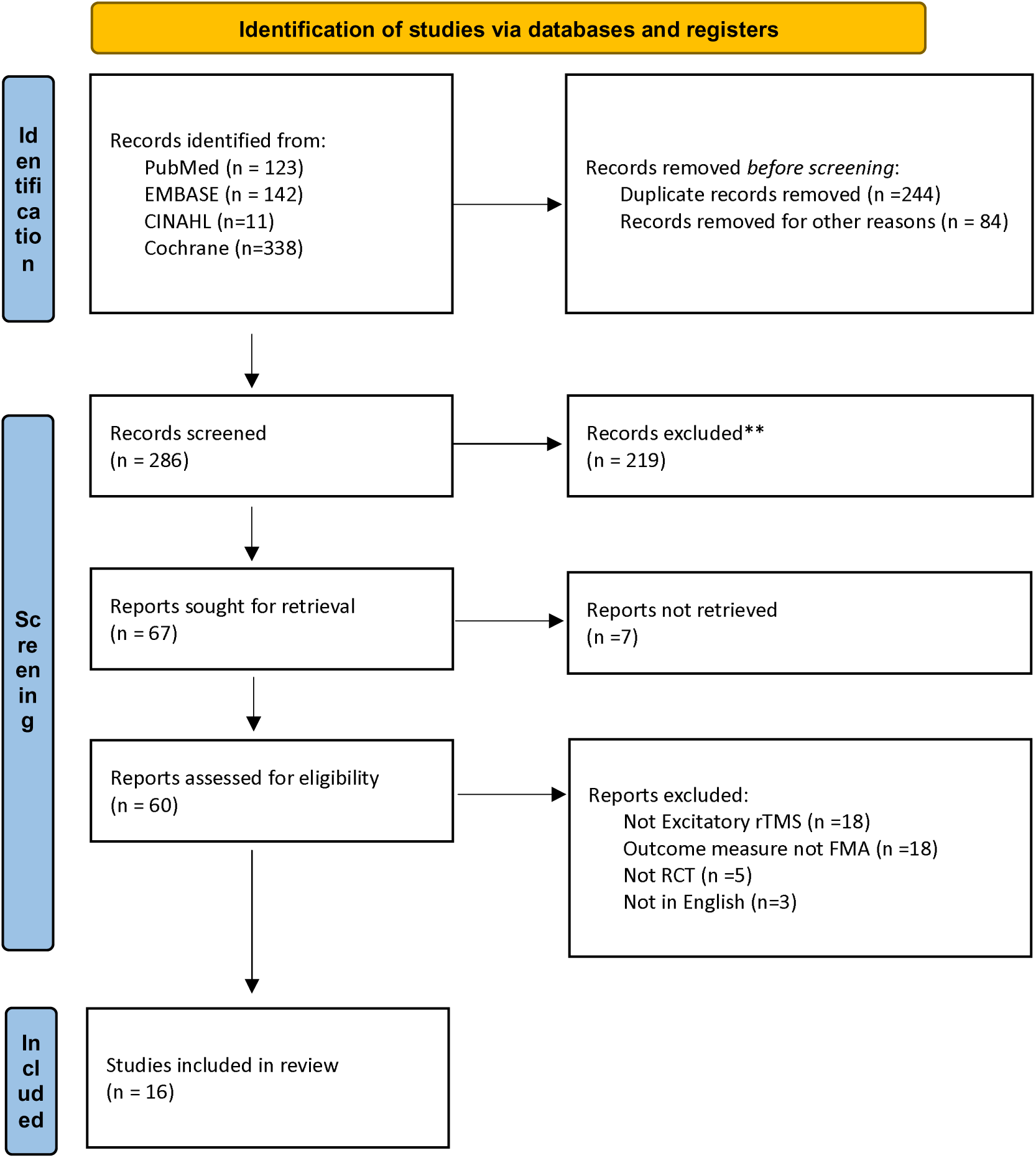
PRISMA Flow diagram depicting retrieval and review process

3.2 Study Characteristics

Study characteristics are described in Table 1. The sample sizes ranged between 12 (Hsu et al., 2013) to 69 (Du et al., 2016) participants, with a cumulative total of 583 stroke patients across the 16 included studies. Ten studies reported adverse events including transient headaches (n=7) (Du et al., 2016; Du et al., 2019; Hsu et al., 2013) and mild-tingling (n=1) (Hsu et al., 2013). An additional 2 patients reported severe adverse effects including epileptic seizures (Chervyakov et al., 2018). However, Chervyakov et al. (2018) acknowledged that they did not utilise any EEG pre-screening protocol prior to commencing rTMS that would have ruled out participants at risk of developing seizures or epileptiform activity. The mean age of patients ranged from 51 (Juan et al., 2022) to 75 years (Watanabe et al., 2018). Three studies included patients more than 6-months after stroke (Ackerley et al., 2016; Chang et al., 2022; Chen et al., 2019), two studies between 5 and 8-months after stroke onset (Chen et al., 2021; Chervyakov et al., 2018), three studies between 1 and 6-months after stroke (Haghighi et al., 2021; Hosomi et al., 2016; Yang et al., 2021) and eight studies within 1-month (Chang et al., 2010; Du et al., 2016; Du et al., 2019; Guan et al., 2017; Hsu et al., 2013; Juan et al., 2022; Ke et al., 2020; Watanabe et al., 2018). The time between stroke onset and the start of rTMS varied from 3 days (Guan et al., 2017) to 20-months (Ackerley et al., 2016). It is worth noting that in an effort to group studies into appropriate SRRR-approved categories, the early-subacute (1-3 months) to late-subacute (3-6 months) phase yielded three articles whose mean time post-stroke averaged roughly 45 (Hosomi et al., 2016), 73 (Yang et al., 2021), and 93 days (Haghighi et al., 2021) respectively. No articles matched our inclusion criteria that addressed patients between 4-6 months post-stroke.

**Table 1.**
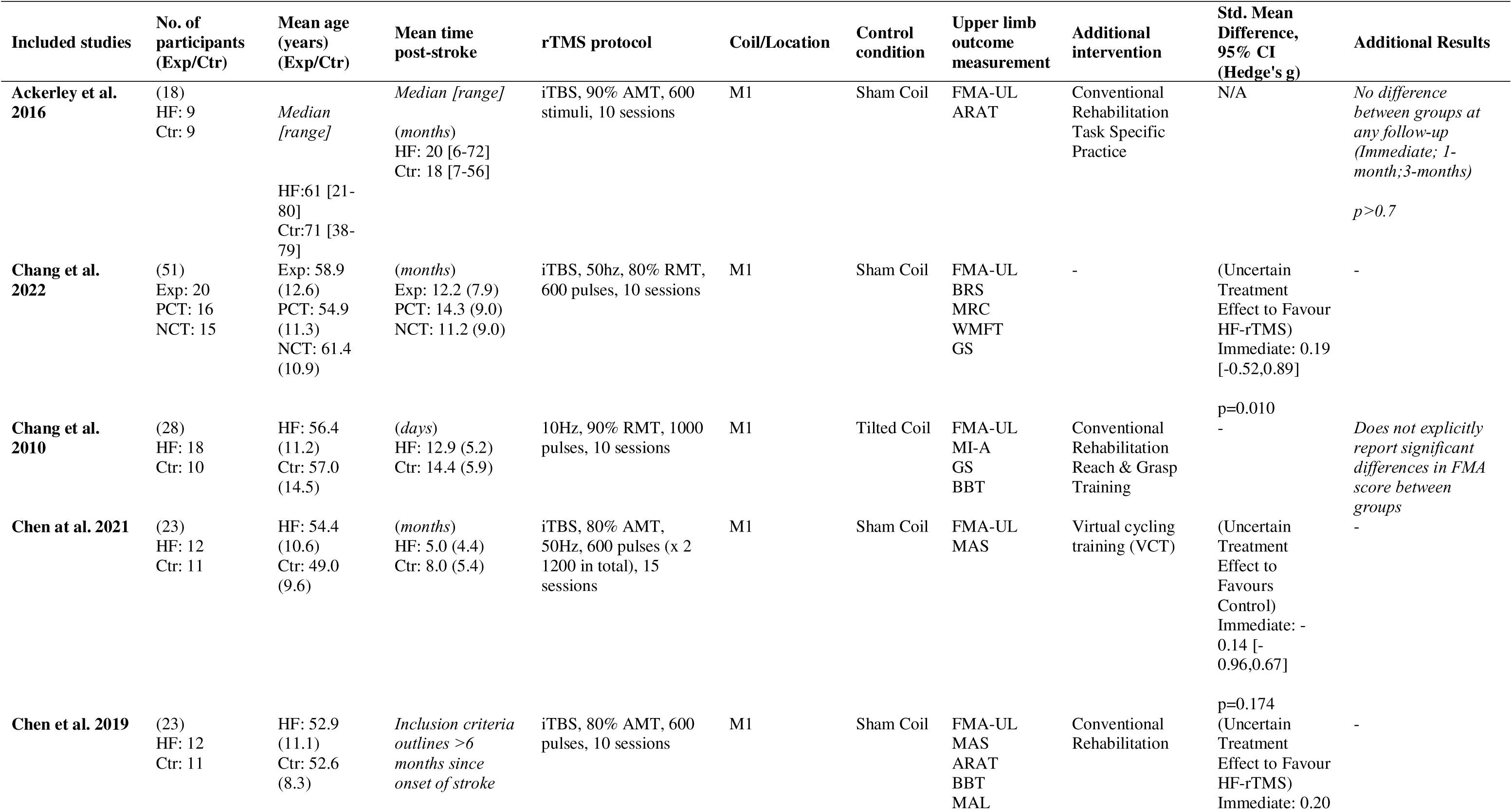
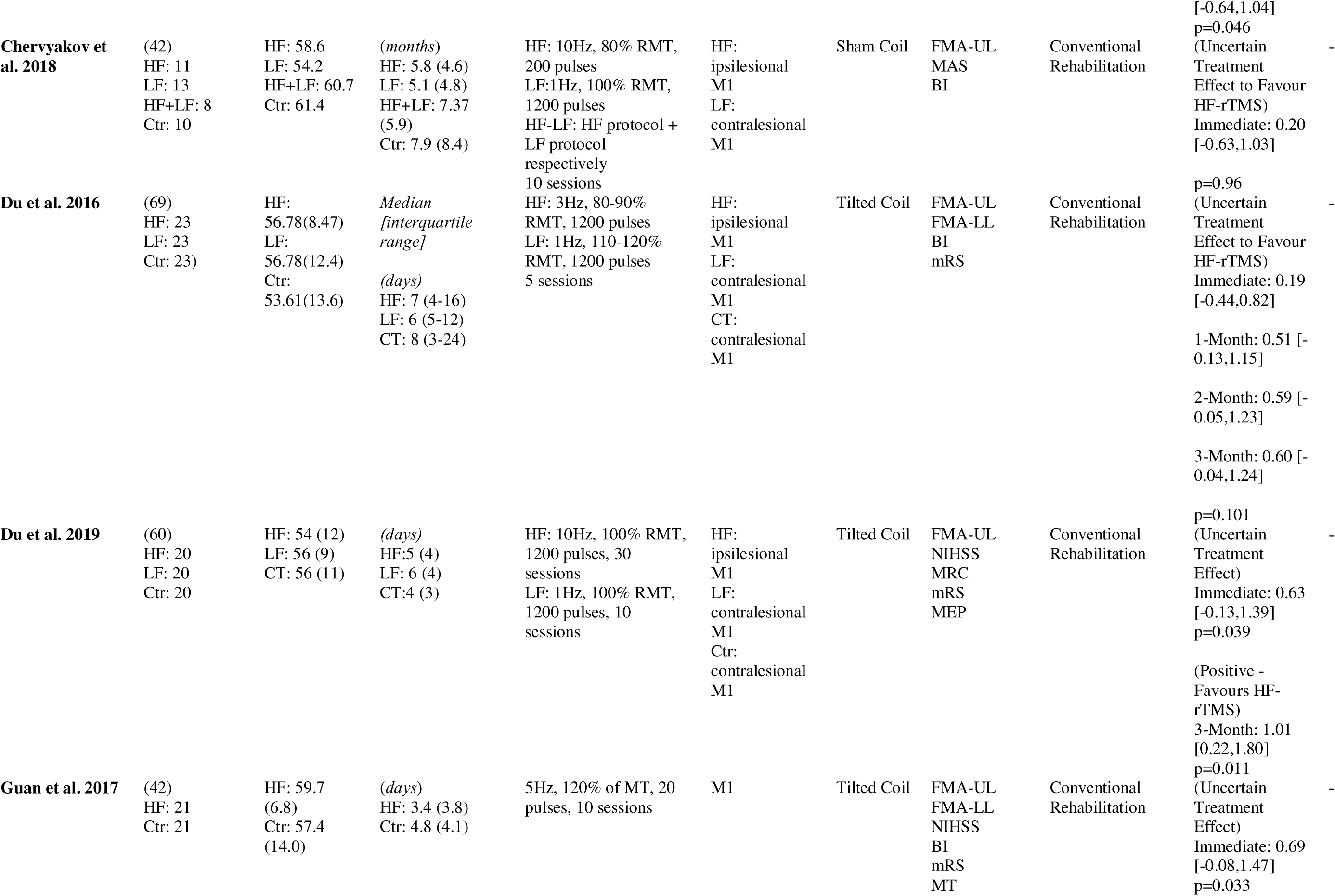
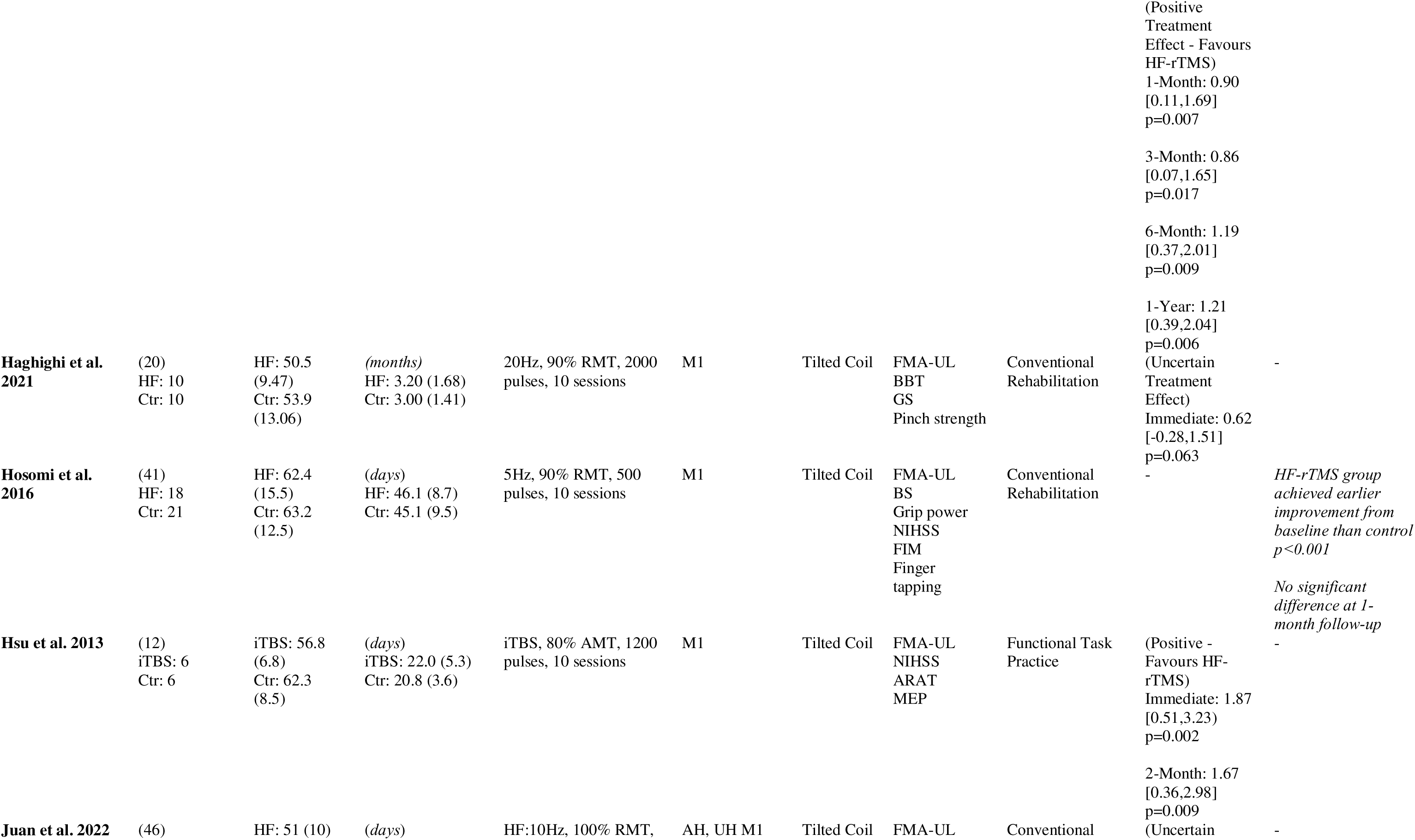
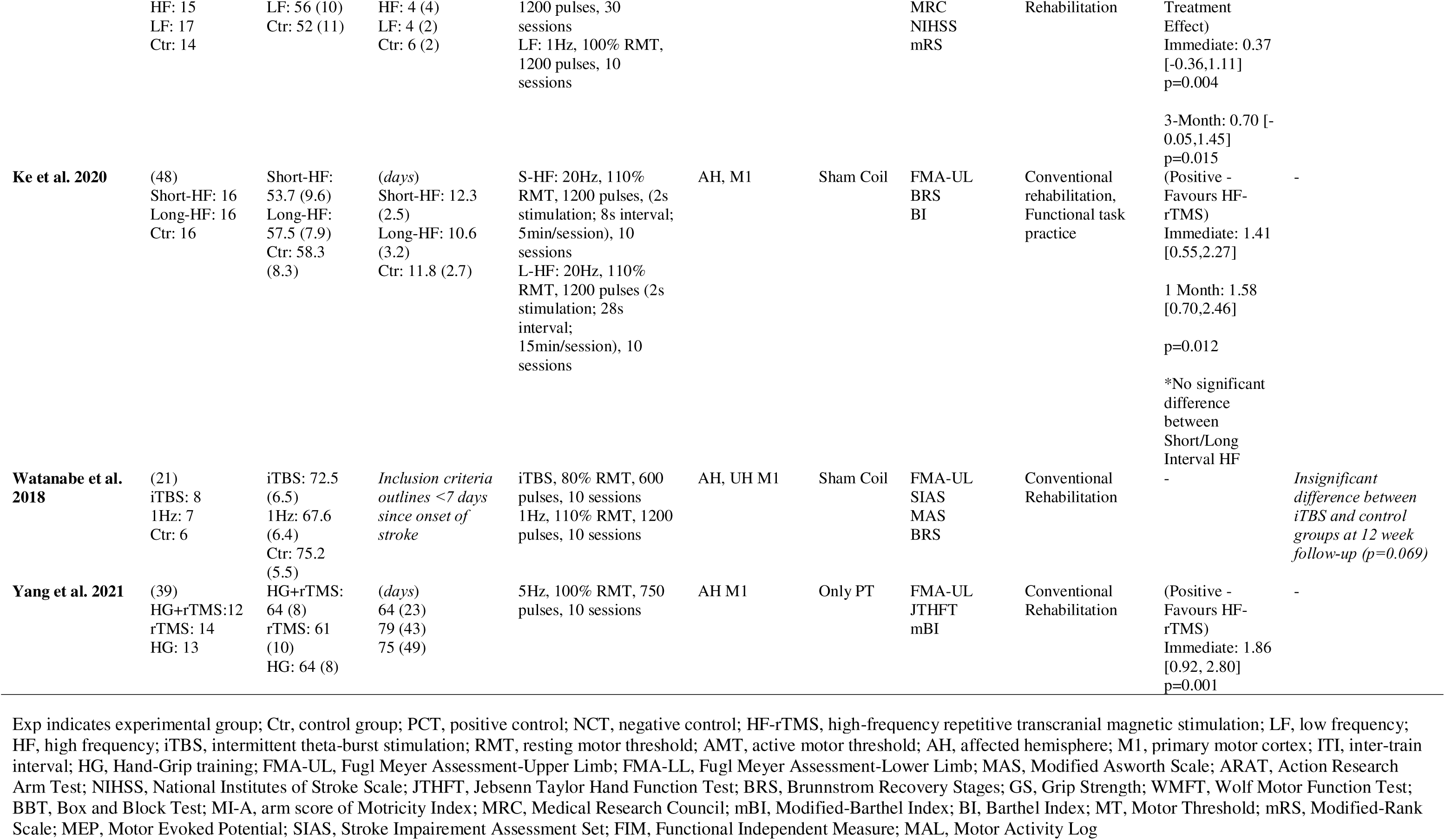
Characteristics and results of included studies.

### 3.3 Treatment Characteristics

The studies included different rTMS treatment protocols, all of which targeted the ipsilesional primary motor cortex with HF-rTMS (see Table 1). The studies used a variety of rTMS frequencies, the most common being 10 Hz (600 – 1200 pulses), used in five studies (Chang et al., 2022; Chervyakov et al., 2018; Du et al., 2019; Guan et al., 2017; Watanabe et al., 2018). Five studies applied a subvariant of rTMS (intermittent theta burst stimulation; iTBS) with two studies applying a TMS intensity of 80% active motor threshold (aMT) for 1200 pulses (Hsu et al., 2013; Watanabe et al., 2018), two studies applying 80% aMT for 600 pulses (Chen et al., 2021; Chen et al., 2019) and one study applying 90% aMT for 600 pulses (Ackerley et al., 2016). Most studies incorporated 10 daily sessions of rTMS or iTBS, with only one study being less frequent (5 sessions) (Du et al., 2016) and two studies more frequent (15 (Chen et al., 2021) and 30 sessions (Du et al., 2019)). All studies used a figure-of-eight coil for real rTMS and iTBS treatment. However, the protocol for the control or comparator group varied between holding the coil perpendicular to the patient’s head (Chang et al., 2010; Du et al., 2016; Du et al., 2019; Guan et al., 2017; Haghighi et al., 2021; Hosomi et al., 2016; Hsu et al., 2013; Juan et al., 2022), flipping the coil (Ackerley et al., 2016; Chang et al., 2022; Chen et al., 2021; Chen et al., 2019; Ke et al., 2020; Watanabe et al., 2018), disconnecting the coil (Chervyakov et al., 2018), or using traditional physical therapy (Yang et al., 2021). Most studies trialled the experimental and control conditions in addition to conventional physical therapy. The experimental groups completed therapy sessions post-rTMS or post-iTBS, with some studies allowing time for consolidation (Ackerley et al., 2016; Chen et al., 2021). One study incorporated 45 mins of virtual cycling training after completing 10 mins upper limb strengthening instead of conventional physical therapy (Chen et al., 2021).

The outcome measures for the upper limb function varied across the studies, with most studies incorporating several different measures. In addition to the Fugl-Meyer Assessment (FMA), studies employed the Action Research Arm Test (ARAT), Brunnstrom stages (BS), Medical Research Council (MRC) Scale, Wolf Motor Function Test, NIH Stroke Scale (NIHSS), Modified Rankin Scale (mRS) and a range of other strength and dexterity assessments (see Table 1). More than half of the studies had post-intervention measurements at multiple time points (n=10), up to 1-year post-stroke (Guan et al., 2017). The remaining studies included outcome measurements immediately post-intervention. Appendix C presents the raw group scores from the FMA extracted from the included studies.

### 3.4 Risk of Bias

Figure 2 summarises the risk of bias assessment results. Two studies (Chang et al., 2010; Haghighi et al., 2021) had some concerns about bias arising from the randomization process due to a lack of information regarding concealing the allocation sequence until patients were enrolled and assigned to the interventions. Three studies (Ke et al., 2020; Watanabe et al., 2018; Yang et al., 2021) had some concerns about bias due to deviations from the intended intervention as there was no information on whether individuals delivering treatment were aware of participants assigned interventions during the trial. Overall, 11 studies were assessed as having low risk of bias (Ackerley et al., 2016; Chang et al., 2022; Chen et al., 2021; Chen et al., 2019; Chervyakov et al., 2018; Du et al., 2016; Du et al., 2019; Guan et al., 2017; Hosomi et al., 2016; Hsu et al., 2013; Juan et al., 2022).

**Figure 2.**
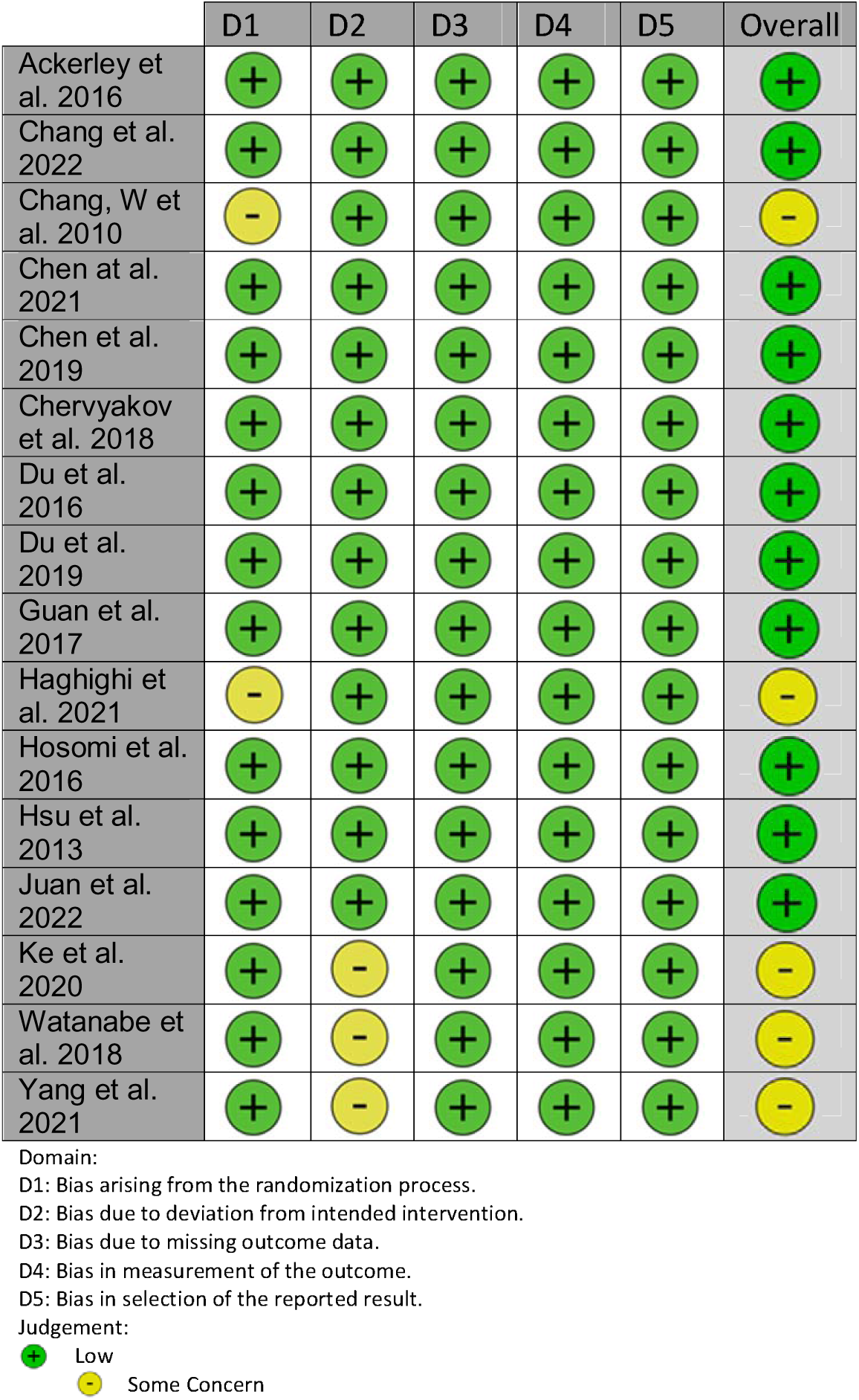
Results from the risk of bias assessment of included studies.

### 3.5 Acute – Early Sub-Acute (<1-month) patients-Assessed Immediately After Intervention

Six studies (Du et al., 2016; Du et al., 2019; Guan et al., 2017; Hsu et al., 2013; Juan et al., 2022; Ke et al., 2020) presented data for participants within 1-month of stroke at the end of their intervention period. Meta-analysis showed a significant effect on FMA-UL score in favour of HF rTMS (mean difference [95% CI] 3.53 [1.82, 5.25]) (see Figure 3A). However, heterogeneity was substantial (*I^2^* = 72%), arising from the high degree of variability in the reported SD results across studies. The GRADE assessment identified a very low level of certainty in a positive effect from HF-rTMS on FMA-UL immediately after intervention in people <1-month post-stroke. All GRADE assessment results are presented in Table 2.

**Figure 3.**
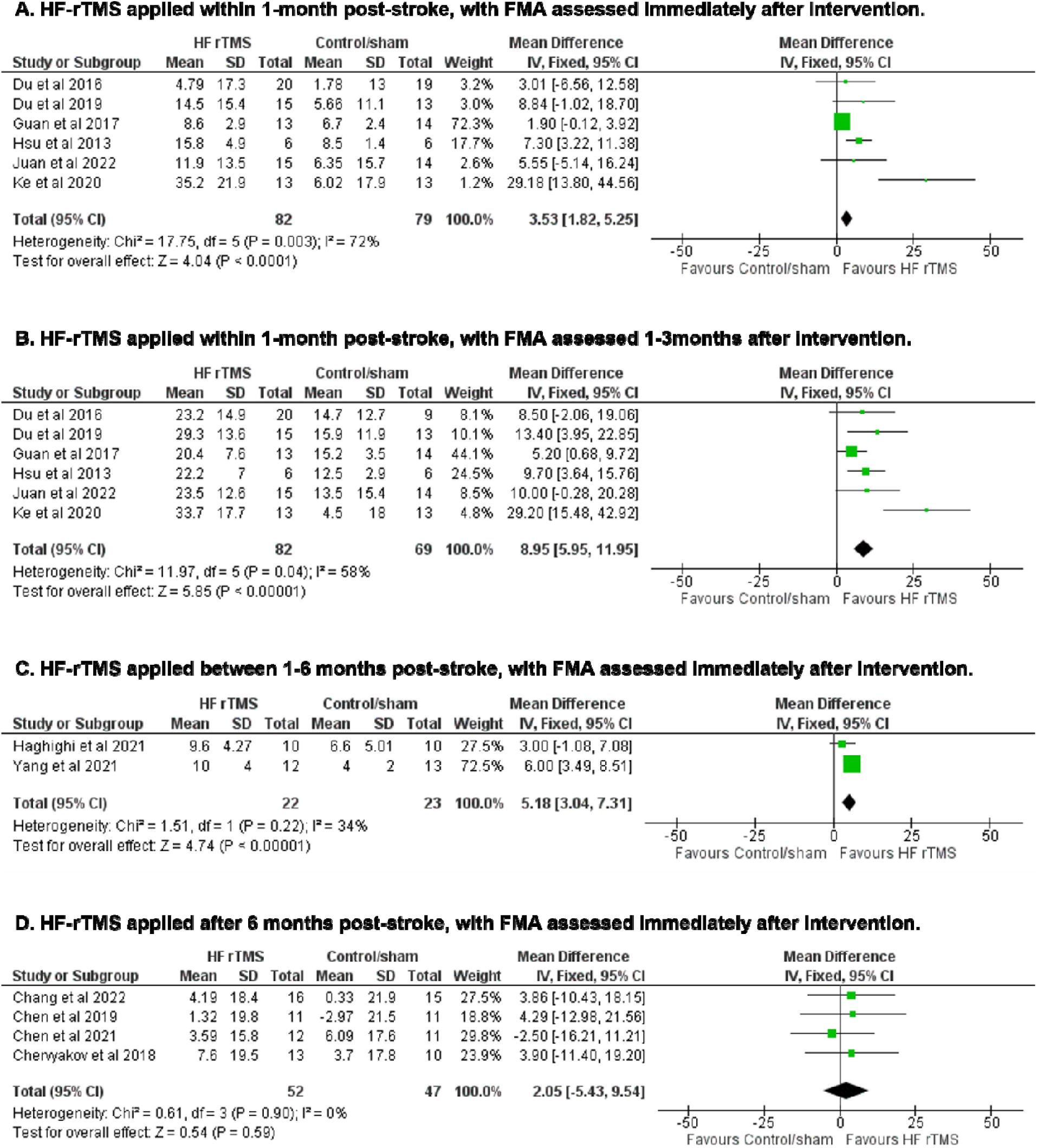
Forest plots displaying results of meta-analyses. The FMA scores in the columns for HF-rTMS and Control/sham represent the mean (SD) change from pre to post-assessment in that group. The mean different (95% CI) column represents the difference (HF-rTMS vs Control/sham) in the change score. **3A.** Effects of HF-rTMS when applied within 1-month post-stroke with FMA assessed immediately after intervention. **3B.** Effects of HF-rTMS when applied within 1-month post-stroke with FMA assessed 1-3 months after intervention. Data were collected at 1 month (Ke et al 2020), 2 months (Hsu et al 2013) and 3 months (Du et al 2016 and 2019; Guan et al 2017; Juan et al 2022) after the end of the intervention. **3C.** Effects of HF-rTMS when applied between 1-6 months post-stroke with FMA assessed immediately after intervention. **3D.** Effects of HF-rTMS when applied after 6-months post-stroke with FMA assessed immediately after intervention.

**Table 2.**
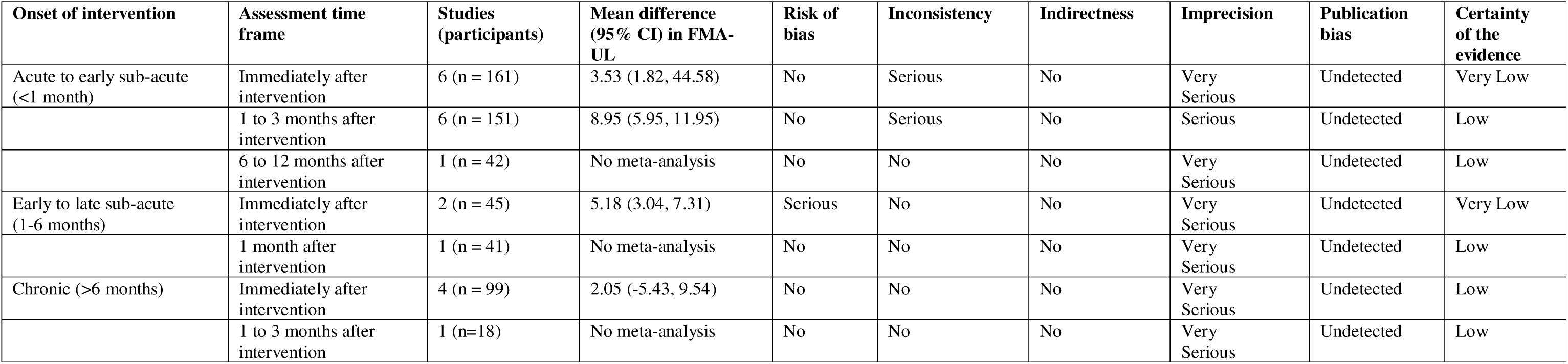
GRADE assessment of the certainty of evidence for upper limb function (FMA-UL) depending on onset of intervention and assessment time frame. A positive mean difference indicates an effect that favours the experimental (rTMS) group.

### 3.6 Acute – Early Sub-Acute (<1-month) patients– Assessed 1-3 Months After Intervention

Six studies (Du et al., 2016; Du et al., 2019; Guan et al., 2017; Hsu et al., 2013; Juan et al., 2022; Ke et al., 2020) presented data for participants within 1-month of stroke, within 1-3 months of the end of their intervention period. Results from the 3-month assessment time point were chosen for Du et al 2016 and Guan et al 2017 over earlier alternatives to avoid study duplication. Meta-analysis showed a significant effect on FMA-UL score in favour of HF rTMS (mean difference [95% CI] 8.95 [5.95, 11.95]) (see Figure 3B). Heterogeneity was moderate to substantial (*I^2^*= 58%). The GRADE assessment identified a low level of certainty in a positive effect from HF rTMS on FMA-UL 1-3 months after intervention in people <1-month post-stroke.

### 3.7 Acute – Early Sub-Acute (<1-month) patients– Assessed 6-12 Months After Intervention

A meta-analysis could not be conducted, as data were only available from one study (Guan et al., 2017). Guan et al. (2017) demonstrated an apparent positive treatment effect which was sustained from 6-months post-stroke (SMD = 1.19, 95% CI 0.37, 2.01) to 12-months post-stroke (SMD = 1.15, 95% CI 0.34, 1.97). The GRADE assessment could be conducted with only one study, with a low level of certainty in a positive effect from HF rTMS on FMA-UL 6-12 months after intervention in people <1-month post-stroke. These results should be interpreted with caution.

### 3.8 Early-to-Late Subacute (1-6 months) patients – Assessed Immediately After Intervention

The effect of rTMS on sub-acute patients was investigated in three studies (Haghighi et al., 2021; Hosomi et al., 2016; Yang et al., 2021). Of these studies, one had insufficient data to be included in the meta-analysis (Hosomi et al., 2016) therefore data from two studies (Haghighi et al., 2021; Yang et al., 2021) for participants 1-6 months after stroke, assessed immediately after the end of their intervention period, were used. Meta-analysis showed a significant effect on FMA-UL score in favour of HF rTMS (mean difference [95% CI] 5.18 [3.04, 7.31]) (see Figure 3C). Heterogeneity was minor (*I^2^* = 34%), as the data from both studies were similar. The GRADE assessment identified a very low level of certainty in a positive effect of HF rTMS on FMA-UL immediately after intervention in people 1-6 months post-stroke.

### 3.9 Early-to-Late Subacute (1-6 months) patients– Assessed 1-Month After Intervention

Hosomi et. al (2016) was the only study that investigated the effects of rTMS on subacute patients compared to sham over multiple periods. The study found a significant within-group improvement in FMA for both intervention and control groups from baseline to days 12 and 29, respectively (both p<0.001). However, there was no difference in FMA scores between groups at 29 days post-stroke (p>0.05). Though these results should be interpreted with caution, the GRADE assessment identified a low level of certainty in the absence of an effect of HF rTMS on FMA-UL one month after intervention in people 1-6 months post-stroke

### 3.10 Chronic (>6-months) patients– Assessed Immediately After Intervention

Four studies (Chang et al., 2022; Chen et al., 2021; Chen et al., 2019; Chervyakov et al., 2018) presented data for participants who were >6-months after stroke, assessed immediately after the end of their intervention period. Meta-analysis showed no significant effect on FMA-UL score from HF rTMS (mean difference [95% CI] 2.05 [-5.45, 9.54]) (see Figure 3D). Heterogeneity was not present (*I^2^* = 0%). The GRADE assessment identified a low level of certainty in the absence of an effect of HF rTMS on FMA-UL immediately after intervention in people >6-months post-stroke.

### 3.11 Chronic (>6-months) patients– Assessed 1-3 Months After Intervention

Ackerley et. al (2016) was the only study that investigated the effects of rTMS on chronic patients compared to sham at both immediate and long term follow-up. The study found no significant between groups differences in FMA-UL scores at any time point (Immediate, 1-Month, and 3-Months) (p>0.05). Though these results should be interpreted with caution, the GRADE assessment identified a low level of certainty in the absence of an effect of HF rTMS on FMA-UL one-to-three months after intervention in people >6-months post-stroke.

## 4. Discussion

The results of this review demonstrated that there is limited yet promising evidence that HF-rTMS leads to greater improvements in FMA-UL score compared with a control group. Despite this, however, the limitations of small sample sizes and varying intervention protocols make definitive conclusions difficult. Unlike that of LF-rTMS, the current body of evidence for the use of HF-rTMS in post-stroke rehabilitation is relatively inconclusive. Prior systematic reviews and meta-analyses confirm the potential utility of HF-rTMS; however, these reviews pool outcome measures that may not reflect the true change in motor function capabilities (Hsu et al., 2012; Vabalaite et al., 2021; van Lieshout et al., 2019; Zhang, Xing, Fan, et al., 2017). The merging of multiple assessment scores amongst individual studies may increase the likelihood of presenting false positives. For this reason, our review focussed solely on the FMA-UL, a more reliable measure of stroke-related upper limb function (Gladstone et al., 2002; van Lieshout et al., 2019). It is important to recognize the factors that may have impacted the effect size between studies. In much of the reported data, those yielding insignificant, or uncertain results commonly presented with sample sizes smaller than n=20 per group (Ackerley et al., 2016; Chang et al., 2022; Chang et al., 2010; Chen et al., 2021; Chen et al., 2019; Chervyakov et al., 2018; Haghighi et al., 2021). As the consensus in the literature suggests that the effects of rTMS may be subtle, particularly in the chronic phase of stroke, much larger sample sizing in future research may be required to produce more clinically relevant results for the effectiveness of HF-rTMS. Lastly, of the varying application protocols (e.g., stimulation frequency, number of pulses delivered, intervals of rTMS application, and the number of sessions involved) in HF-rTMS within our review, there were no identifiable trends in particular parameters that yielded more or less significant results between studies. However, formal meta-analyses should be conducted to determine possible optimal stimulation parameters to improve UL function following stroke using HF rTMS.

### 4.1 Timing of rTMS Treatment Post-Stroke

#### 4.1.1 Acute to Early-Subacute (< 1 m)

In the acute phase of stroke, current literature suggests a positive effect of HF-rTMS on FMA-UL scores immediately following intervention. While the studies included in this review showed a significant effect in favour of HF-rTMS, results were of low to very low certainty. This is likely due to small sample sizing with considerable variances in scores. However, a majority of studies that conducted follow-up assessments between 1-3 months post-intervention reported evidence of lasting favourable effects of HF-rTMS in acute stroke patients with significantly less degree of heterogeneity (I^2^=58% vs 72%), compared to immediate assessment (Chang et al., 2010; Du et al., 2016; Du et al., 2019; Guan et al., 2017; Hsu et al., 2013; Juan et al., 2022; Ke et al., 2020). These results are consistent with previous literature suggesting that recovery of motor function may be enhanced with the application of rTMS within the first-month post-stroke (Hsu et al., 2012; Vabalaite et al., 2021; van Lieshout et al., 2019; Zhang, Xing, Fan, et al., 2017). The evidence within our review suggests that the benefits of HF-rTMS on acute-stroke patients become more noticeable and enduring over time. While findings indicate a significant improvement difference between groups when assessed immediately after intervention, subsequent follow-up data implies that sustained and more pronounced effects can appear from 1-3 months post-intervention ((Du et al., 2016; Du et al., 2019; Guan et al., 2017; Hsu et al., 2013; Juan et al., 2022; Ke et al., 2020). Amongst our review, Guan et al. (2017) was the only study that conducted long-term follow-up assessments at the 6- and 12-month mark on patients assessed and treated with HF-rTMS in the acute-to-early subacute stroke phase. Their findings reveal a significant and much larger effect at both time points (6, and 12-months) in favour of HF-rTMS. This suggests long-lasting effects of treatment, compared to conventional therapy. However, the certainty of the results varies, and the lack of comparable study parameters highlights the need for further investigation to clarify the long-term efficacy of HF-rTMS. Overall, the favourable results exhibited in the acute to early-subacute phase are consistent with the literature regarding the neurophysiological principles of stroke rehabilitation (Alia et al., 2017; Kleim & Jones, 2008; Zeiler & Krakauer, 2013). It is well established that the first-month post-stroke has the greatest capacity for neuroplastic change, and is expected to demonstrate the most significant and reliable improvements in motor function. Conventional rehabilitation in the first-month post-stroke may be enhanced with the synergistic application of HF-rTMS. However, additional research should look to replicate these findings with larger samples to draw more conclusive evidence.

#### 4.1.2. Early-Subacute to Late-Subacute (1m-6m)

The articles included in this review conveyed positive evidence of the effectiveness of HF-rTMS on immediate post-intervention assessment in the early-to late-subacute phase of stroke. Although Haghighi et al. (2021) reported insufficient findings (95% CI [-1.08, 7.08]), their results illustrate a trend towards significance. This lack of certainty is likely attributed to their relatively low sample size of 10 participants per group. In contrast, Yang et al. (2021) showed a significant improvement in FMA-UL scores in individuals treated with HF-rTMS compared to those exposed to the sham/control. While the heterogeneity between both studies was minor (I^2^=34%), the lack of additional comparator studies makes it challenging to draw definitive conclusions. Furthermore, Hosomi et al. (2016) examined an additional assessment at one month; however, preliminary results indicate no differences between groups. Further research is recommended to provide a conclusive statement on the potentially lasting effects of HF-rTMS administered in the subacute phases of stroke. Currently, there is a lack of high-quality evidence regarding rehabilitation at this stage (García-Rudolph et al., 2019; Hsu et al., 2012). Recent publications have suggested that although the timing-dependent effectiveness of HF-rTMS in the subacute phase may not be as favourable as in the acute phase, it may still be more beneficial when compared to the chronic phase (Hsu et al., 2012; van Lieshout et al., 2019; Zhang, Xing, Fan, et al., 2017).

#### 4.1.3 Chronic (>6-months)

The compilation of evidence in this review demonstrates that there is an uncertain and inconclusive effect as to the immediate effect on motor function performance with the application of ipsilesional HF-rTMS during the chronic phase of stroke. Discrepancies between individual study findings may be attributed to the rTMS application protocol as well as varying sample sizes, though heterogeneity was not present between studies (I^2^=0%). In addition to rTMS parameters, the inclusion of functional task-specific training, as opposed to conventional rehabilitation, may have affected outcomes (French et al., 2016; Hatem et al., 2016). It is important to consider that Chervyakov et al. (2018) used a different classification system to justify patients within the chronic stroke phase, as suggested by previous researchers (Adeyemo et al., 2012; Bahn et al., 1996). While the average time from onset of stroke was over 6-months, the inclusion range in Chervyakov et al. (2018) was between 6-weeks and 12-months post-stroke. Their findings should therefore be considered accordingly due to their wide range of applicable candidates. Similar to the results illustrated in the immediate assessment findings in patients in the chronic stroke phase, Ackerley et al. (2016) determined that the application of HF-rTMS produced a negligible effect on FMA-UL scores 1-3 months post-intervention when compared to the sham/control group. However, more studies investigating the potential long-term effects of HF-rTMS treatment in individuals in the chronic stroke phase are needed to produce more conclusive evidence.

The results concluded in this review are consistent with recent systematic and meta-analyses, which report insufficient evidence for the application of rTMS during the chronic phase of stroke (Hsu et al., 2012; van Lieshout et al., 2019; Zhang, Xing, Fan, et al., 2017). The evidence in the literature illustrates that HF-rTMS in conjunction with conventional rehabilitation beginning 6-months post-stroke and beyond has minimal capacity to elicit significant neurophysiological change (Hsu et al., 2012; van Lieshout et al., 2019). This decrease in magnitude for effective rehabilitation is likely due to a plateau effect in synaptic plasticity (Cramer, 2008; Kwakkel et al., 2017).

### 4.2 Comparison to Previous Studies

The trend towards a positive effect of HF-rTMS on upper limb motor function in stroke patients is in agreement with the majority of the current literature. A meta-analysis conducted by Vabalaite and colleagues (2021) found that HF-rTMS may improve upper limb motor function better than sham stimulation. Two meta-analyses looked at both low and high-frequency rTMS, and although the latter was found to have a positive effect on upper limb motor function, the contralesional LF-rTMS was more effective (Hsu et al., 2012; Zhang, Xing, Fan, et al., 2017). Both studies support the concept of interhemispheric differences in excitability being altered in stroke and contributing to impaired motor function (Hsu et al., 2012). In addition to comparing rTMS frequencies, both studies assessed rTMS at different time points post-stroke, finding increased effects in the acute to subacute phase (0 to 6-months) compared to chronic (>6-months) (Hsu et al., 2012; van Lieshout et al., 2019; Zhang, Xing, Fan, et al., 2017). It must be noted, however, that these two studies pooled outcome measures together, potentially overestimating the effect size. Another potentially confounding factor in previous meta-analyses when making conclusions between LF- and HF-rTMS was the disparity between the total number of studies that assessed either. A vast majority of studies included in reviews investigated the use of LF-rTMS, making it difficult to make accurate comparisons between the two methods.

### 4.3 Strengths and Limitations

By restricting studies to only those which included the FMA as an outcome measure, the likelihood of detecting a true change in upper limb motor function was increased. As outcome measures at the level of function are closely linked to stroke-related neural changes it was concluded that this would be more sensitive to motor recovery (van Lieshout et al., 2019). Additionally, by isolating one outcome measure as opposed to several, the likelihood of false positives was reduced. A thorough search of multiple databases was used and a robust quality assessment tool was applied to evaluate any included RCTs. By subcategorizing the data into stroke phases and follow-up assessment time points, the ability to discern trends regarding stroke phases and any long-lasting effects of HF-rTMS was enhanced. Appropriate statistical measures were taken to account for variables that may overestimate effect size. Correctional factors were used to address small sample sizing between studies. All calculations and data extraction processes were based on the Cochrane Handbook for Systematic Reviews of Intervention to ensure methodologically appropriate analysis of data.

The present study contains various limitations to be addressed. First and foremost, small sample sizes within the included studies led to a large degree of uncertainty in subsequent results. Secondly, due to limited data, we could not account for differences in rTMS protocols, namely intensities and duration. Thirdly, the methodological quality of studies was assessed using the ROB2 protocol, while this is a robust assessment, it is reliant on a critical appraisal conducted by the authors and as such is subject to human error. Interrater reliability was found to be low between 2 of the 4 reviewers, for a small proportion of total full-text reviews. This may have implications regarding errors introduced into the study, however, this is likely to be minimal as the remaining combination of reviewers had near-perfect agreeableness for the majority of the full-text article (McHugh, 2012). Lastly, there was significant heterogeneity in the application protocol and the timing of assessments, which led to difficulty in drawing firm comparisons.

### 4.4 Implications for Practice

HF-rTMS applied to the ipsilesional cortex may provide some short and long-lasting effects on motor recovery post-stroke in the acute-to-subacute phase, however, current evidence is conflicting and further research is required. Furthermore, benefits are accentuated when applied early post-stroke (<1m) during the window of increased neuroplasticity. Improvements due to rTMS are more likely to be detected with outcome measures aimed at the function level (i.e., FMA), which may be an important consideration in the clinical and research setting (Kwakkel et al., 2017; van Lieshout et al., 2019). The combination of HF-rTMS and high doses of task-specific physical therapy aimed at correcting post-stroke functional deficits may yield greater improvements in function (Hayward & Brauer, 2015). The use of HF-rTMS has been demonstrated to be safe with no significant adverse events as a result of stimulation reported in the studies. It is known that rTMS can result in transient side effects such as headache, local pain and hearing changes, although these are uncommon. Seizures are perhaps the most concerning potential side-effect of rTMS, however, this can be mitigated through adequate screening and risk evaluation (Chervyakov et al., 2018).

### 4.5 Conclusion

While the use of HF-rTMS on the ipsilesional M1 undoubtedly shows promise in terms of functional upper limb motor recovery post-stroke, more research into its efficacy is warranted. Similarly, there is tentative evidence to suggest a long-lasting effect duration of HF-rTMS, but the strength of this evidence is weak. Current research is highly heterogeneous, particularly regarding outcome measures and follow up assessments. These disparities combined with small sample sizes have introduced a high degree of uncertainty to the results. Larger sample sizes and the development of a standardised set of measurements to assess upper limb function may be an appropriate means of combating this. Finally, future research should include follow up measurements at varying time points post-intervention, including both a 6 and 12-month follow up to enable a proper understanding of any long-lasting effects.

## Author Contributions

Nazzareno Russo: Conceptualisation, Investigation, Formal analysis, Writing – Original Draft, Writing – Review & Editing

Thomas Hunt: Conceptualisation, Investigation, Formal analysis, Writing – Original Draft, Writing – Review & Editing

Alex McMullin: Conceptualisation, Investigation, Formal analysis, Writing – Original Draft, Writing – Review & Editing

Timothy Payard: Conceptualisation, Investigation, Formal analysis, Writing – Original Draft, Writing – Review & Editing

Elise Gane: Conceptualisation, Methodology, Formal analysis, Writing – Review & Editing

Martin Sale: Conceptualisation, Writing – Review & Editing, Supervision

## Data Availability

All data produced in the present work are contained in the manuscript.

## Acknowledgements

This review was conducted within a university-based course, and the authors wish to acknowledge the assistance received from the tutor, Dr Atiyeh Vaezipour.

## Appendix A. Database Search Strategy

### Pubmed

(((((((((((((((((((((((((((Stroke[MeSH Terms]) OR (Brain Ischemia[MeSH Terms])) OR (Intracranial Hemorrhage[MeSH Terms])) OR (stroke[Title/Abstract]))) OR (“cerebrovascular accident“[Title/Abstract])) OR (CVA[Title/Abstract])) OR (“cerebrovascular ischemia“[Title/Abstract])) OR (“cerebrovascular ischaemia“[Title/Abstract])) OR (“brain ischemia“[Title/Abstract])) OR (“brain ischaemia“[Title/Abstract])) OR (“ischemic stroke“[Title/Abstract])) OR (“haemorrhagic stroke“[Title/Abstract])) OR (brain infarction[Title/Abstract])) OR (“intracerebral hemorrhage“[Title/Abstract])) OR (“intracerebral haemorrhage“[Title/Abstract])) OR (intracranial thromb*[Title/Abstract])) OR (“lacunar infarct“[Title/Abstract])) OR (“lacunar stroke“[Title/Abstract])) OR (poststroke[Title/Abstract])) OR (“post stroke“[Title/Abstract])) OR (“brain hemorrhage“[Title/Abstract])) OR (“brain haemorrhage“[Title/Abstract])) OR (“cerebrovascular accident“[Text Word])) OR (CVA[Text Word])) OR (“ischemic stroke“[Text Word])) OR (“haemorrhagic stroke“[Text Word])) OR (“hemorrhagic stroke“[Text Word])

#### AND

(((((transcranial magnetic stimulation[MeSH Terms]) OR (transcranial magnetic stimulation[Title/Abstract])) OR (transcranial magnetic stimulation[Text Word])) OR (transcranial magnetic stimulation, repetitive[MeSH Terms])) OR (repetitive transcranial magnetic stimulation[Title/Abstract])) OR (repetitive transcranial magnetic stimulation[Text Word])

#### AND

(((((((((((((((Upper Extremity[MeSH Terms]) OR (upper extremity*[Title/Abstract])) OR (arm[Title/Abstract])) OR (arms[Title/Abstract])) OR (hand[Title/Abstract])) OR (hands[Title/Abstract])) OR (Elbow[MeSH Terms])) OR (Wrist[MeSH Terms])) OR (Shoulder[MeSH Terms])) OR (forearm[Title/Abstract])) OR (finger*[Title/Abstract])) OR (elbow[Title/Abstract])) OR (wrist[Title/Abstract])) OR (shoulder[Title/Abstract])) OR (finger*[Text Word])) OR (forearm[Text Word])

#### AND

((((((((((((((muscle strength[MeSH Terms]) OR (psychomotor performance[MeSH Terms])) OR (motor skills[MeSH Terms])) OR (proprioception[MeSH Terms])) OR (activities of daily living[MeSH Terms])) OR (stroke rehabilitation[MeSH Terms])) OR (exercise movement techniques[MeSH Terms])) OR (sensation[MeSH Terms])) OR (coordination[Title/Abstract])) OR (coordination[Text Word])) OR (dexterity[Title/Abstract])) OR (dexterity[Text Word])) OR (muscle spasticity[MeSH Terms])) OR (“muscle strength“[Text Word])) OR (“upper limb function“[Title/Abstract])

### CINAHL

( ( (MH “Stroke, Lacunar”) OR (MH “Embolic Stroke”) OR (MH “Stroke Patients”) OR (MH “Hemorrhagic Stroke”) OR (MH “Ischemic Stroke+”) OR (MH “Stroke+”) ) OR ( MH ( stroke or cerebrovascular accident or cva or cerebrovascular event or cve or transient ischaemic attack or tia or intracranial hemorrhage or intracranial haemorrhage or post stroke or poststroke or cerebral infarct or intracerebral haemorrhage or intracerebral hemorrhage or lacunar infarct) ) ) OR TI ( ( stroke or cerebrovascular accident or cva or cerebrovascular event or cve or transient ischaemic attack or tia or intracranial hemorrhage or intracranial haemorrhage or post stroke or poststroke or cerebral infarct or intracerebral haemorrhage or intracerebral hemorrhage or lacunar infarct) ) OR AB ( ( stroke or cerebrovascular accident or cva or cerebrovascular event or cve or transient ischaemic attack or tia or intracranial hemorrhage or intracranial haemorrhage or post stroke or poststroke or cerebral infarct or intracerebral haemorrhage or intracerebral hemorrhage or lacunar infarct) )

#### AND

MH ( transcranial magnetic stimulation or tms or repetitive transcranial stimulation or rtms ) OR TI ( transcranial magnetic stimulation or tms or repetitive transcranial magnetic stimulation or rtms ) OR AB ( transcranial magnetic stimulation or tms or repetitive transcranial magnetic stimulation or rtms )

#### AND

MH ( upper extremity or upper limb or hand or arm or wrist or elbow or finger or forearm or shoulder ) OR TI ( upper extremity or upper limb or hand or arm or wrist or elbow or finger or forearm or shoulder ) OR AB ( upper extremity or upper limb or hand or arm or wrist or elbow or finger or forearm or shoulder )

#### AND

MH ( upper limb function or arm function or hand function or upper extremity function or strength or dexterity or coordination or proprioception or sensation or psychomotor skills or “activities of daily living” or muscle spasticity or motor skills ) OR TI ( upper limb function or arm function or hand function or upper extremity function or strength or dexterity or coordination or proprioception or sensation or psychomotor skills or “activities of daily living” or muscle spasticity or motor skills ) OR AB ( upper limb function or arm function or hand function or upper extremity function or strength or dexterity or coordination or proprioception or sensation or psychomotor skills or “activities of daily living” or muscle spasticity or motor skills )

### Embase

(’stroke’/exp OR ‘stroke’ OR ‘brain ischemia’/exp OR ‘brain ischemia’ OR ‘intracranial hemorrhage’/exp OR ‘intracranial hemorrhage’ OR ‘stroke’:ab,ti OR ‘cerebrovascular accident’:ab,ti OR ‘cva’:ab,ti OR ‘cerebrovascular ischemia’:ab,it OR ‘brain ischemia’:ab,ti OR ‘ischemic stroke’:ab,ti OR ‘hemorrhagic stroke’:ab,ti OR ‘brain infarction’:ab,ti OR ‘intracerebral hemorrhage’:ab,ti OR ‘intracranial thromb’:ab,ti OR ‘lacunar infarct’:ab,ti OR ‘lacunar stroke’:ab,ti OR ‘poststroke’:ab,ti OR ‘post stroke’:ab,ti OR ‘brain hemorrhage’:ab,ti OR ‘brain haemorrhage’:ab,ti OR ‘cerebrovascular accident’/exp OR ‘cerebrovascular accident’ OR ‘cva’/exp OR ‘cva’ OR ‘ischemic stroke’/exp OR ‘ischemic stroke’ OR ‘haemorrhagic stroke’/exp OR ‘haemorrhagic stroke’ OR ‘hemorrhagic stroke’/exp OR ‘hemorrhagic stroke’) AND [embase]/lim AND [randomized controlled trial]/lim AND [english]/lim

#### AND

(’transcranial magnetic stimulation’/exp OR ‘transcranial magnetic stimulation’:ab,ti OR ‘transcranial magnetic stimulation’ OR ‘transcranial magnetic stimulation, repetitive’/exp OR ‘transcranial magnetic stimulation, repetitive’:ab,ti OR ‘repetitive transcranial magnetic stimulation’:ab,ti OR ‘repetitive magnetic stimulation’) AND [embase]/lim AND [randomized controlled trial]/lim AND [english]/lim

#### AND

(’upper limb’/exp OR ‘upper limb’:ab,ti OR ‘upper extremity’/exp OR ‘upper extremity’:ab,ti OR ‘arm’:ab,ti OR ‘arms’:ab,ti OR ‘shoulder’/exp OR ‘shoulder’:ab,ti OR ‘elbow’/exp OR ‘elbow’:ab,ti OR ‘hand’/exp OR ‘hand’:ab,ti OR ‘finger’/exp OR ‘finger’:ab,ti OR ‘finger’ OR ‘hand’ OR ‘shoulder’ OR ‘elbow’ OR ‘forearm’ OR ‘forearm’:ab,ti) AND [embase]/lim AND [randomized controlled trial]/lim AND [english]/lim

#### AND

(’muscle strength’/exp OR ‘psychomotor performance’/exp OR ‘motor skills’/exp OR ‘proprioception’/exp OR ‘activities of daily living’/exp OR ‘stroke rehabilitation’/exp OR ‘exercise movement techniques’/exp OR ‘sensation’/exp OR ‘coordination’:ab,ti OR ‘coordination’ OR ‘dexterity’:ab,ti OR ‘dexterity’ OR ‘muscle spasticity’/exp OR ‘muscle strength’ OR ‘upper limb function’:ab,ti) AND [embase]/lim AND [randomized controlled trial]/lim AND [english]/lim

### Cochrane

Stroke [MeSH descriptor] OR Cerebral Infarction [MeSH descriptor] OR Cerebral Hemorrhage [MeSH descriptor] OR Stroke, Lacunar [MeSH descriptor] OR (stroke OR “cerebrovascular accident” OR CVA OR “ischemic stroke” OR “haemorrhagic stroke” OR “brain infarction” OR “lacunar stroke”):ti,ab,kw OR (stroke OR “cerebrovascular accident” OR CVA OR “ischemic stroke” OR “haemorrhagic stroke” OR “brain infarction” OR “lacunar stroke”)

#### AND

Transcranial Magnetic Stimulation [MeSH descriptor] OR (“transcranial magnetic stimulation” OR “repetitive transcranial magnetic stimulation”):ti,ab,kw OR (“transcranial magnetic stimulation” OR “repetitive transcranial magnetic stimulation” OR TMS OR rTMS)

#### AND

Upper Extremity [MeSH descriptor] OR (“upper extremity” OR arm OR hand OR elbow OR wrist OR shoulder OR forearm OR finger):ti,ab,kw OR (“upper extremity” OR arm OR hand OR elbow OR wrist OR shoulder OR forearm OR finger)

#### AND

Muscle Strength [MeSH descriptor] OR Psychomotor Performance [MeSH descriptor] OR Motor Skills [MeSH descriptor] OR Proprioception [MeSH descriptor] OR Stroke Rehabilitation [MeSH descriptor] OR Sensation OR (“upper extremity function” OR “upper limb function” OR “hand function” OR strength OR “muscle strength” OR dexterity OR coordination OR proprioception OR sensation OR psychomotor skills OR motor skills OR “stroke rehabilitation” OR “exercise movement techniques”):ti,ab,kw OR (“upper extremity function” OR “upper limb function” OR “hand function” OR strength OR “muscle strength” OR dexterity OR coordination OR proprioception OR sensation OR psychomotor skills OR motor skills OR “stroke rehabilitation” OR “exercise movement techniques”)

## Appendix B. Interrater reliability measured using Cohen’s Kappa

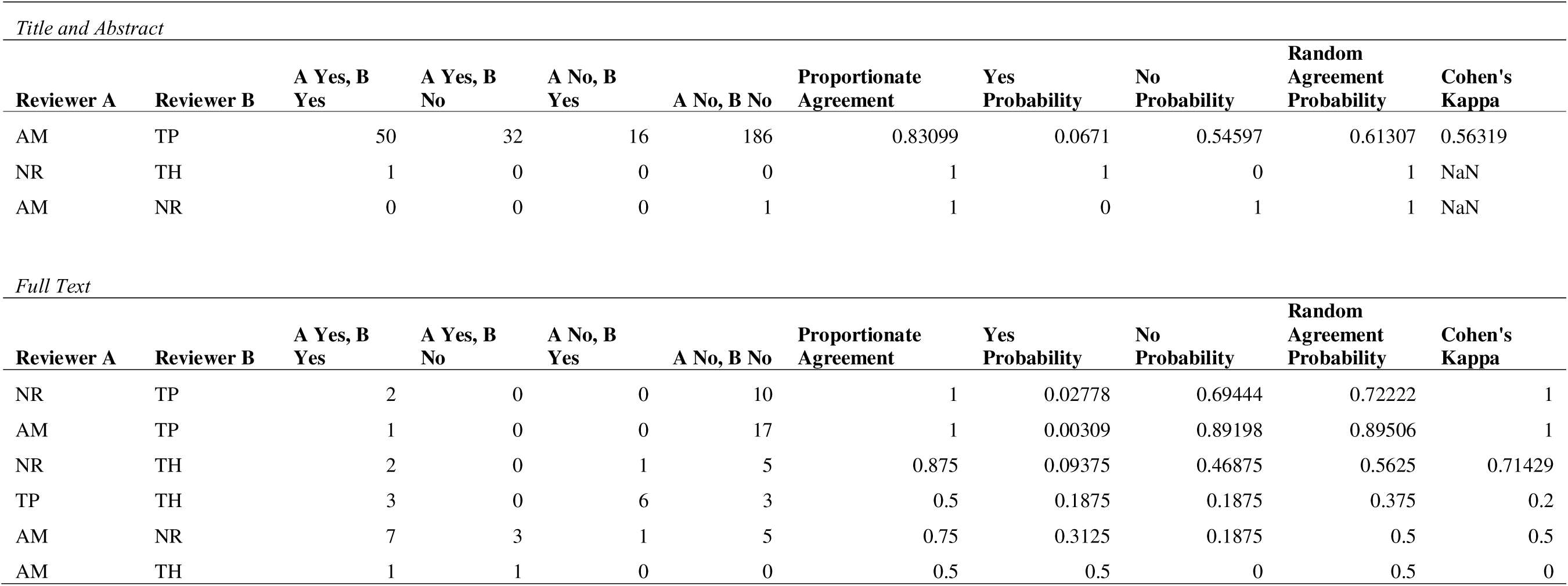

## Appendix C. Raw data from included studies. Studies by Ackerley et al (2016), Chang W et al (2010), Hosomi et al (2016) and Watanabe et al (2018) did not have suitable group-level data for extraction

| Study | Pre-program HF rTMS | | Pre-program Control/Sham | | Post-program assessment | Post-program HF rTMS | | Post-program Control/Sham | | Between-group difference ΔHF-ΔControl |
| --- | --- | --- | --- | --- | --- | --- | --- | --- | --- | --- |
|  | Mean (SD) | N | Mean (SD) | N |  | Mean (SD) | N | Mean (SD) | N | Mean difference (95% CI) |
| Chang et al. 2022 | 82.50 (18.8) | 16 | 85.67(21.85) | 15 | Immediate | 85.95(17.85) | 16 | 86.00(21.95) | 15 | 3.86 [-10.43, 18.15] |
| Chen at al. 2021 | 43.58(15.35) | 12 | 34.55(18.34) | 11 | Immediate | 47.17(16.30) | 12 | 40.64(16.83) | 11 | -2.50[-16.21, 11.21] |
| Chen et al. 2019 | 33.33(19.80) | 11 | 30.03(22.11) | 11 | Immediate | 34.65(19.80) | 11 | 27.06(20.79) | 11 | 4.29[-12.98, 21.56] |
| Chervyakov et al. 2018 | 33.3(18.2) | 13 | 32.3(15.8) | 10 | Immediate | 40.7(20.65) | 13 | 36.0(19.51) | 10 | 3.90[-11.40,19.20] |
| Du et al. 2016 | 25.91(17.17) | 20 | 22.52(13.28) | 19 | Immediate | 30.70(17.52) | 20 | 24.30(12.79) | 19 | 3.01[-6.56,12.58] |
|  |  |  |  |  | 1 month | 39.13(16.48) | 20 | 27.98(12.08) | 19 | 7.76[4.66,10.86] |
|  |  |  |  |  | 2 months | 44.44(14.18) | 20 | 32.41(12.13) | 19 | 8.64[5.66,11.62] |
|  |  |  |  |  | 3 months | 49.08(12.26) | 20 | 37.22(12.03) | 19 | 8.47[5.59,11.35] |
| Du et al. 2019 | 29.00(16.00) | 15 | 21.9(11.13) | 13 | Immediate | 43.48(14.87) | 15 | 27.56(10.97) | 13 | 8.82[4.87,12.77] |
|  |  |  |  |  | 3 months | 58.3(10.75) | 15 | 37.84(12.56) | 13 | 13.36[9.60,17.12] |
| Guan et al. 2017 | 37.40(9.80) | 13 | 40.9(8.9) | 14 | Immediate | 45.9(9.1) | 13 | 47.7(8.1) | 14 | 1.7[-1.05,4.45] |
|  |  |  |  |  | 1 month | 51.1(6.8) | 13 | 53.8(6.0) | 14 | 0.8[-1.65,3.25] |
|  |  |  |  |  | 3 months | 57.5(3.7) | 13 | 59.4(3.6) | 14 | 1.6[-0.57,3.77] |
|  |  |  |  |  | 6 months | 59.2(1.8) | 13 | 60.8(2.3) | 14 | 2.7[0.64,4.76] |
|  |  |  |  |  | 1 year | 60.0(2.1) | 13 | 61.5(1.8) | 14 | 2[0.07,4.07] |
| Haghighi et al. 2021 | 30.50(6.78) | 10 | 36.7(7.71) | 10 | Immediate | 40.10(9.49) | 10 | 43.3(10.28) | 10 | 3.00[-1.08,7.08] |
| Hsu et al. 2013 | 35.8(11.8) | 6 | 37.0(12.4) | 6 | Immediate | +15.7(4.8)<sup>1</sup> | 6 | +8.5(1.4)<sup>1</sup> | 6 | N.R.<sup>1</sup> |
|  |  |  |  |  | 2 months | +22.2(9.0)<sup>1</sup> | 6 | +12.5(2.9)<sup>1</sup> | 6 | N.R.<sup>1</sup> |
| Juan et al. 2022 | 33.0(14.0) | 15 | 22.39(14.27) | 14 | Immediate | 44.92(13.01) | 15 | 28.74(16.96) | 14 | 5.55[-5.14-16.24] |
|  |  |  |  |  | 3 months | 56.5(10.99) | 15 | 35.86(16.43) | 14 | 10.03[6.05,14.01] |
| Ke et al. 2020 | 34.5(12.4) |  | 41.5(20.5) |  | Immediate | 69.7(28.39) | 13 | 47.52(14.83) | 13 | 29.18[13.8,44.56] |
|  |  |  |  |  | 1 month | 68.22(21.79) | 13 | 46.04(15.11) | 13 | 29.18[23.51,34.85] |
| Yang et al. 2021 | 47(6) | 12 | 47(7) | 13 | Immediate | 57(5) | 12 | 51(7) | 13 | 6.00[3.49,8.51] |

Footnotes: <sup>1</sup>Article presented only baseline and within-group change from baseline scores. N.R. = not reported.

### PRISMA 2020 item checklist

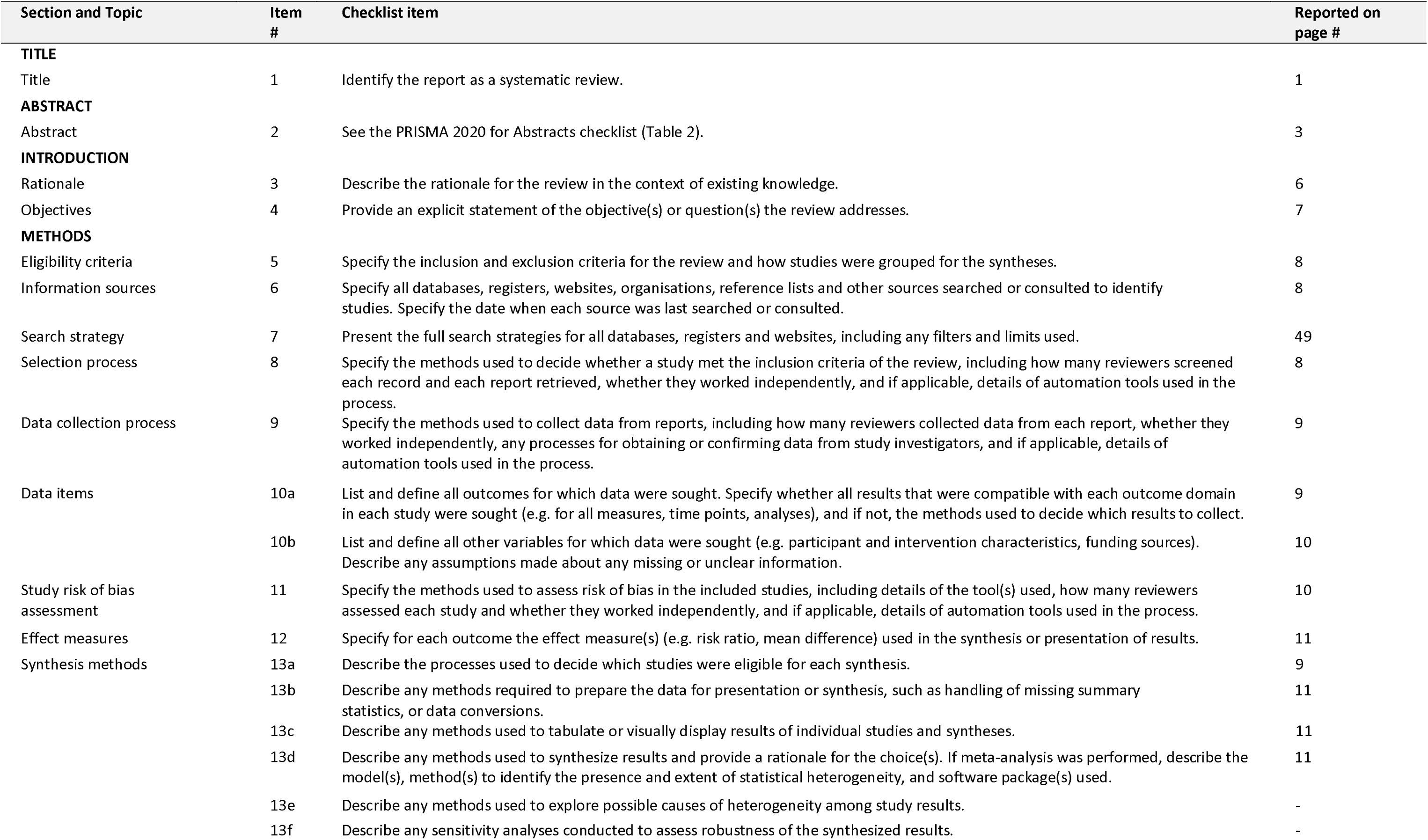
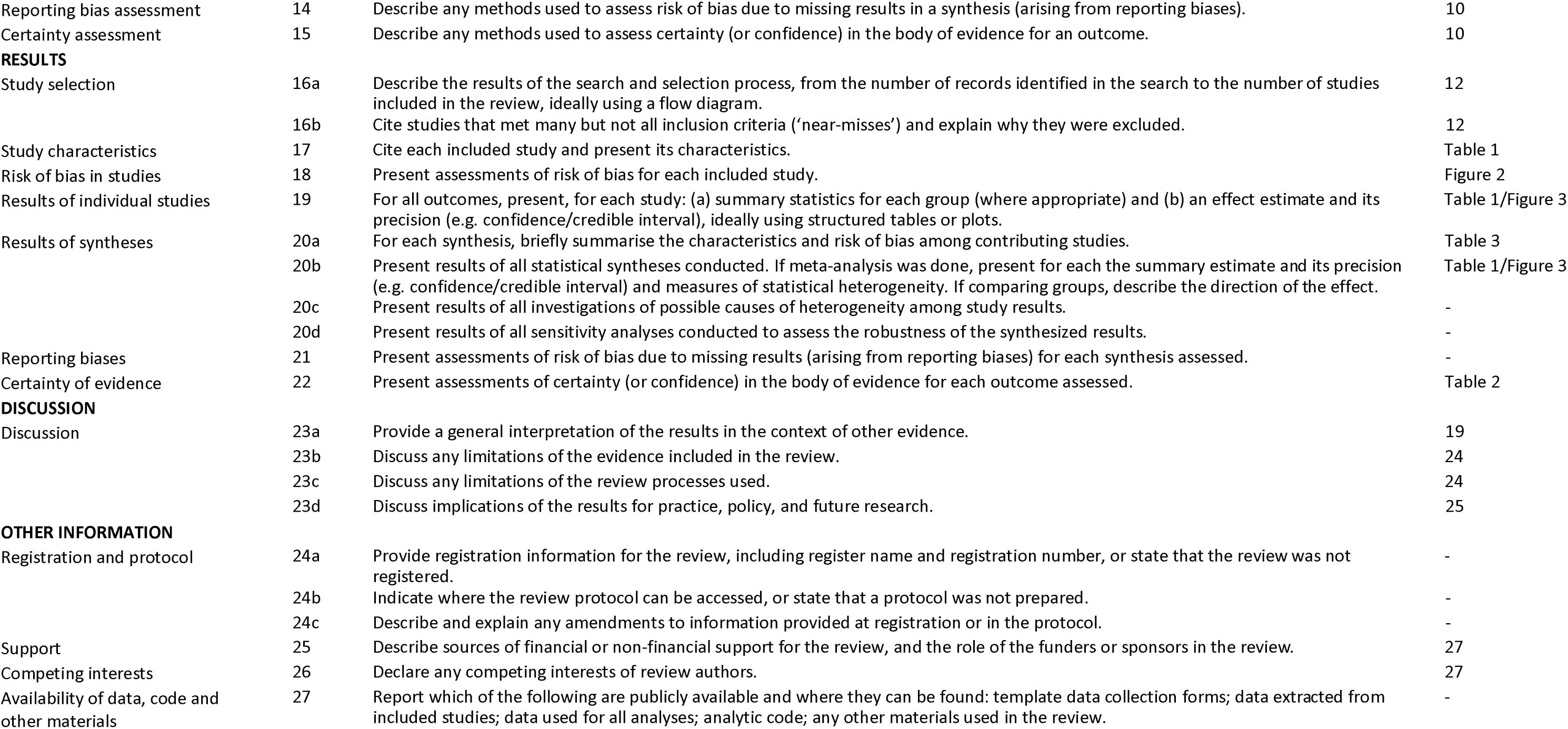

| Section and Topic | Item # | Checklist item | Reported on page # |
| --- | --- | --- | --- |
| **TITLE** |  |  |  |
| Title | 1 | Identify the report as a systematic review. | 1 |
| **ABSTRACT** |  |  |  |
| Abstract | 2 | See the PRISMA 2020 for Abstracts checklist (Table 2). | 3 |
| **INTRODUCTION** |  |  |  |
| Rationale | 3 | Describe the rationale for the review in the context of existing knowledge. | 6 |
| Objectives | 4 | Provide an explicit statement of the objective(s) or question(s) the review addresses. | 7 |
| **METHODS** |  |  |  |
| Eligibility criteria | 5 | Specify the inclusion and exclusion criteria for the review and how studies were grouped for the syntheses. | 8 |
| Information sources | 6 | Specify all databases, registers, websites, organisations, reference lists and other sources searched or consulted to identify studies. Specify the date when each source was last searched or consulted. | 8 |
| Search strategy | 7 | Present the full search strategies for all databases, registers and websites, including any filters and limits used. | 49 |
| Selection process | 8 | Specify the methods used to decide whether a study met the inclusion criteria of the review, including how many reviewers screened each record and each report retrieved, whether they worked independently, and if applicable, details of automation tools used in the process. | 8 |
| Data collection process | 9 | Specify the methods used to collect data from reports, including how many reviewers collected data from each report, whether they worked independently, any processes for obtaining or confirming data from study investigators, and if applicable, details of automation tools used in the process. | 9 |
| Data items | 10a | List and define all outcomes for which data were sought. Specify whether all results that were compatible with each outcome domain in each study were sought (e.g. for all measures, time points, analyses), and if not, the methods used to decide which results to collect. | 9 |
|  | 10b | List and define all other variables for which data were sought (e.g. participant and intervention characteristics, funding sources). Describe any assumptions made about any missing or unclear information. | 10 |
| Study risk of bias assessment | 11 | Specify the methods used to assess risk of bias in the included studies, including details of the tool(s) used, how many reviewers assessed each study and whether they worked independently, and if applicable, details of automation tools used in the process. | 10 |
| Effect measures | 12 | Specify for each outcome the effect measure(s) (e.g. risk ratio, mean difference) used in the synthesis or presentation of results. | 11 |
| Synthesis methods | 13a | Describe the processes used to decide which studies were eligible for each synthesis. | 9 |
|  | 13b | Describe any methods required to prepare the data for presentation or synthesis, such as handling of missing summary statistics, or data conversions. | 11 |
|  | 13c | Describe any methods used to tabulate or visually display results of individual studies and syntheses. | 11 |
|  | 13d | Describe any methods used to synthesize results and provide a rationale for the choice(s). If meta-analysis was performed, describe the model(s), method(s) to identify the presence and extent of statistical heterogeneity, and software package(s) used. | 11 |
|  | 13e | Describe any methods used to explore possible causes of heterogeneity among study results. | - |
|  | 13f | Describe any sensitivity analyses conducted to assess robustness of the synthesized results. | - |
| Reporting bias assessment | 14 | Describe any methods used to assess risk of bias due to missing results in a synthesis (arising from reporting biases). | 10 |
| Certainty assessment | 15 | Describe any methods used to assess certainty (or confidence) in the body of evidence for an outcome. | 10 |
| **RESULTS** |  |  |  |
| Study selection | 16a | Describe the results of the search and selection process, from the number of records identified in the search to the number of studies included in the review, ideally using a flow diagram. | 12 |
|  | 16b | Cite studies that met many but not all inclusion criteria ('near-misses') and explain why they were excluded. | 12 |
| Study characteristics | 17 | Cite each included study and present its characteristics. | Table 1 |
| Risk of bias in studies | 18 | Present assessments of risk of bias for each included study. | Figure 2 |
| Results of individual studies | 19 | For all outcomes, present, for each study: (a) summary statistics for each group (where appropriate) and (b) an effect estimate and its precision (e.g. confidence/credible interval), ideally using structured tables or plots. | Table 1/Figure 3 |
| Results of syntheses | 20a | For each synthesis, briefly summarise the characteristics and risk of bias among contributing studies. | Table 3 |
|  | 20b | Present results of all statistical syntheses conducted. If meta-analysis was done, present for each the summary estimate and its precision (e.g. confidence/credible interval) and measures of statistical heterogeneity. If comparing groups, describe the direction of the effect. | Table 1/Figure 3 |
|  | 20c | Present results of all investigations of possible causes of heterogeneity among study results. | - |
|  | 20d | Present results of all sensitivity analyses conducted to assess the robustness of the synthesized results. | - |
| Reporting biases | 21 | Present assessments of risk of bias due to missing results (arising from reporting biases) for each synthesis assessed. | - |
| Certainty of evidence | 22 | Present assessments of certainty (or confidence) in the body of evidence for each outcome assessed. | Table 2 |
| **DISCUSSION** |  |  |  |
| Discussion | 23a | Provide a general interpretation of the results in the context of other evidence. | 19 |
|  | 23b | Discuss any limitations of the evidence included in the review. | 24 |
|  | 23c | Discuss any limitations of the review processes used. | 24 |
|  | 23d | Discuss implications of the results for practice, policy, and future research. | 25 |
| **OTHER INFORMATION** |  |  |  |
| Registration and protocol | 24a | Provide registration information for the review, including register name and registration number, or state that the review was not registered. | - |
|  | 24b | Indicate where the review protocol can be accessed, or state that a protocol was not prepared. | - |
|  | 24c | Describe and explain any amendments to information provided at registration or in the protocol. | - |
| Support | 25 | Describe sources of financial or non-financial support for the review, and the role of the funders or sponsors in the review. | 27 |
| Competing interests | 26 | Declare any competing interests of review authors. | 27 |
| Availability of data, code and other materials | 27 | Report which of the following are publicly available and where they can be found: template data collection forms; data extracted from included studies; data used for all analyses; analytic code; any other materials used in the review. | - |

### PRISMA 2020 for Abstracts checklist*

**PRISMA 2020 for Abstracts checklist\***
| Section and Topic | Item # | Checklist item | Completed |
| --- | --- | --- | --- |
| **TITLE** |  |  |  |
| Title | 1 | Identify the report as a systematic review. | Y |
| **BACKGROUND** |  |  |  |
| Objectives | 2 | Provide an explicit statement of the main objective(s) or question(s) the review addresses. | Y |
| **METHODS** |  |  |  |
| Eligibility criteria | 3 | Specify the inclusion and exclusion criteria for the review. | N |
| Information sources | 4 | Specify the information sources (e.g. databases, registers) used to identify studies and the date when each was last searched. | Y |
| Risk of bias | 5 | Specify the methods used to assess risk of bias in the included studies. | Y |
| Synthesis of results | 6 | Specify the methods used to present and synthesize results. | Y |
| **RESULTS** |  |  |  |
| Included studies | 7 | Give the total number of included studies and participants and summarise relevant characteristics of studies. | Y |
| Synthesis of results | 8 | Present results for main outcomes, preferably indicating the number of included studies and participants for each. If meta-analysis was done, report the summary estimate and confidence/credible interval. If comparing groups, indicate the direction of the effect (i.e. which group is favoured). | Y |
| **DISCUSSION** |  |  |  |
| Limitations of evidence | 9 | Provide a brief summary of the limitations of the evidence included in the review (e.g. study risk of bias, inconsistency and imprecision). | Y |
| Interpretation | 10 | Provide a general interpretation of the results and important implications. | Y |
| **OTHER** |  |  |  |
| Funding | 11 | Specify the primary source of funding for the review. | N |
| Registration | 12 | Provide the register name and registration number. | N |

\*This abstract checklist retains the same items as those included in the PRISMA for Abstracts statement published in 2013 (48), but has been revised to make the wording consistent with the PRISMA 2020 statement and includes a new item recommending authors specify the methods used to present and synthesize results (item #6)

